# Limited evidence of spill over of antimicrobial resistant *Klebsiella pneumoniae* from animal/environmental reservoirs to humans in India

**DOI:** 10.1101/2024.03.09.24303758

**Authors:** Jobin John Jacob, Aravind V, Benjamin S. Beresford-Jones, Binesh Lal Y, Chaitra Shankar, Yesudoss M, Fiza Abdullah, Monisha Priya T, Sanika Kulkarni, Stephen Baker, Balaji Veeraraghavan, Kamini Walia

## Abstract

**Background:** *Klebsiella pneumoniae* is a common opportunistic pathogen known for having virulent and antimicrobial resistance (AMR) phenotypes. In addition to accumulating AMR and virulence genes, *K. pneumoniae* serves as a vehicle for broadly disseminating these elements into other species. Here, we applied genomic surveillance in a one-health framework to assess the impact of the human-animal-environment interface on AMR transmission.

**Methods:** We sequenced representative genomes of *Klebsiella pneumoniae* isolated from clinical specimens (*n=59*), livestock samples (*n=71*), and hospital sewage samples (*n=16*) from a two-year surveillance study. We compared the taxonomic and genomic distribution of *K. pneumoniae*, the abundance of AMR, virulence genes, and mobile genetic elements between isolates from three sources.

**Results:** *Klebsiella* spp. was the second most commonly isolated species (*n=2,569*). The clonal distribution of *K. pneumoniae* suggested isolates originating from livestock were clonally distinct from those derived from clinical/hospital effluent isolates. Clinical and hospital sewage isolates typically carried a higher number of resistance/virulence genes. There was limited overlap of *K. pneumoniae* clones, AMR genes, virulence determinants, and plasmids between the different settings.

**Conclusion:** Currently, the spread of XDR or hypervirulent clones of *K. pneumoniae* appears to be confined to humans with no clear linkage with non-clinical sources. Moreover, emerging convergent clones of *K. pneumoniae* carrying both resistance and virulence determinants (ST231, ST2096) are likely to have emerged in hospital settings rather than in animal or natural environments. These data challenge the current view of AMR transmission in *K. pneumoniae* in a One-Health context.

## Background

*Klebsiella pneumoniae* are the causative agents of different types of healthcare-associated infections including pneumonia, bloodstream infections, liver abscesses, urinary tract infections, etc.^1, 2^ Multi-drug resistant (MDR) and extremely-drug resistant (XDR) *K. pneumoniae* infections have been rising in hospitals worldwide.^3^ In addition, carbapenem-resistant *K. pneumoniae* (CR-Kp), a particularly hard-to-treat phenotype of the bacteria, has become a public health concern.^4, 5^ In light of this, the World Health Organization (WHO) has listed extended-spectrum β-lactam (ESBL)-producing and CR *K. pneumoniae*as a pathogen of critical priority.^6^ In India alone, *K. pneumoniae* accounts for 18% of all bloodstream infections, and 57% of these organisms are resistant to carbapenems.^7^

Besides being an important nosocomial pathogen, *K. pneumoniae* are widely distributed in the environmental niches and are capable of colonizing and infecting a broad variety of hosts.^8^ The ability of the organism to infect a wide variety of hosts has been attributed to the high plasticity of the accessory genome, which allows them to acquire AMR genes and virulence factors via horizontal gene transfer.^9^ It is therefore not surprising that many of the new AMR genes were identified primarily in *K. pneumoniae* before spreading to other bacterial pathogens.^8^ Therefore a ‘One Health’ perspective on AMR transmission has been postulated, where environmental niches and livestock are considered as potential sources and routes for resistance gene transmission.^10^ Considering the importance of environmental niches in the persistence and spread of AMR genes, it is important to investigate any possibility of such an event occurring in *K. pneumoniae*.^8^ Although multiple studies have indicated the role of animals and the environment in the transmission of genetic determinants associated with AMR into clinical settings conclusive evidence is still lacking.^11, 12^ Conversely, recent findings suggest limited spread of antimicrobial resistance (AMR) genes within the context of One Health, especially in the case of *K. pneumoniae*.^13, 14^

An integrated approach of whole genome sequencing (WGS) and phylogenetic analysis has proven to be an effective tool in determining AMR transmission pathways.^15, 16, 17, 18^ One-Health genomic surveillance studies have previously been used to gain a better understanding of AMR transmission at the human-animal-environment interface.^10, 19^ Here, we used comparative genome analysis of clinical isolates of *K. pneumoniae* from a tertiary-care teaching hospital with non-clinical isolates collected from livestock and wastewater samples from Vellore in Southern India.

## Methods

### Ethical Statement

No human subjects were directly involved in the study. The data of clinical isolates was obtained from the electronic laboratory register of Department of Clinical Microbiology, Christian Medical College, Vellore. All the patient identifying variables in the data were removed before the analysis. Institutional Review Board (IRB) of Christian Medical College (CMC), Vellore, India gave ethical approval for this work vide IRB Min no. 12626 dated 26.02.2020. As the required data had been collected as part of the standard of care for diagnosis, informed consent waiver was granted by the IRB committee CMC, Vellore. The Declaration of Helsinki norms were adhered to while conducting the study.

### Study Area

The study was conducted in the Vellore corporation area located in the Vellore district on the northern side of the Tamil Nadu state in India (Suppl Figure 1). The Vellore corporation has been divided into four zones, and our samples are primarily sourced from the Vellore fort and Katpadi areas (Suppl Table 2).

### Clinical isolates

This study utilized a subset of clinical isolates of *K. pneumoniae* that were isolated from blood cultures at the Christian Medical College (CMC), Vellore between 2020-2022. CMC, Vellore is a 2,234-bed quaternary care hospital with an annual volume of ∼68,000 blood cultures located in Vellore, Southern India.

### Specimen Collection

Animal specimens were collected from farms, slaughterhouses and retail meat shops as per the directive 2003/99/EC of the European Parliament. All samples were placed in sterile plastic containers, stored at 4°C, and processed within 24 hours of the time of sample collection. Biosafety clearance and Ethical committee clearance for this study are obtained from the Institutional Research Board and Ethics Committee of the Institution (IRB Min. No. 12626 dated: 26/02/2020).

Hospital effluents from CMC Vellore sewage plant (12.9258, 79.1343), were collected as per the Biomedical Waste Management Rules, 2016 ^20^ (Suppl Table 3). All samples were stored at 4°C and processed within 24=h of sample collection.

### Sample processing

Clinical Samples: Clinical isolates confirmed as *Klebsiella* sp. from patients diagnosed with bacteremia and pneumonia between 2020 - 2022 were collected at the Department of Clinical Microbiology, Christian Medical College, Vellore. During the study period, 2398 isolates of *Klebsiella* sp. were recovered from blood and sputum samples. Isolates were sub-cultured on MacConkey agar for antibiotic susceptibility testing and DNA extraction.

Sewage Samples: A total of 45 specimens were collected from hospital sewage sites. An aliquot of 100 mL of raw hospital effluent and 200 mL of treated effluent was centrifuged at 10000 rpm for 10 min; the pellet was dissolved in 5 mL of sterile phosphate buffer saline (PBS). These were incubated for 18-24h at 37°C.

Livestock Samples: A total of 229 samples were collected from Livestock. About 2ml of Sterile Todd Hewitt Broth (THB) was added to the meat/intestine specimens and incubated overnight at 37°C and then sub-cultured onto MacConkey agar plates. These were incubated for 18-24h at 37°C. In addition to standard culture, biochemical, and identification procedures, MALDI TOF MS (VITEK® MS, bioMérieux) was also used for speciation.

### Antimicrobial susceptibility testing

Antimicrobial susceptibility testing to cefepime, ceftazidime, piperacillin-tazobactam, meropenem, amikacin, gentamicin and minocycline was performed by Kirby Bauer disc diffusion method and results were interpreted according to CLSI 2020, 2021 and 2022 guidelines. Colistin susceptibility was tested using broth microdilution for the isolates from hospital effluent and animal meat specimens and interpreted according to EUCAST 2020 ^21^ and guidelines published by the Clinical and Laboratory Standards Institute.^22^

### Whole Genome sequencing and Genome analysis

A subset of 146 isolates (Clinical-59; Livestock-71; Hospital sewage-16) characterized as *Klebsiella* sp. were subjected to WGS. DNA was extracted from the pelleted cells using Wizard DNA purification kit (Promega) as per the manufacturer’s protocol.

Sequencing ready, paired-end library was prepared using 100ng of DNA with the Nextera DNA sample preparation kit as per the manufacturer’s instructions (Illumina, Inc., San Diego, USA). Sequencing was then performed (2 X 200 bp) on the Illumina HiSeq platform (MedGenome Labs, Bengaluru). Adapters and indexes were removed from raw reads by using MultiQC (https://github.com/ewels/MultiQC). The filtered high-quality reads were assembled using SPAdes (https://github.com/ablab/spades). Genome annotations were performed using Bakta annotation pipeline v.1.6.8 (https://github.com/oschwengers/bakta).

Antimicrobial resistance genes and plasmids were obtained from the assembled genomes using Abricate (https://github.com/tseemann/abricate) utilizing databases available at NCBI (AMRFinderPlus v.3.11.3) (https://github.com/ncbi/amr) and PlasmidFinder database (https://bitbucket.org/genomicepidemiology/plasmidfinder/src/master/). MLST, virulence genes, resistance and virulence scores were obtained using Kleborate (https://github.com/katholt/Kleborate) and VFDB, Virulence Factor Database (http://www.mgc.ac.cn/VFs/).

### Genome data acquisition and Curation

For comparison, *K. pneumoniae* genomes available at the public databases (NCBI SRA; https://www.ncbi.nlm.nih.gov/pathogens/) sourced from India. Assembled genomes consisting of more than 150 contigs were removed and error-free sequences with higher sequencing depth were only included in the study. A total of 185 genomes sourced from different geographical regions in India were used as the representative collection.

### Minimum Spanning Tree

The assembled *Klebsiella* genomes were searched for core genes based on the Ridom SeqSphere^+^ cgMLST scheme (https://www.ridom.de/seqsphere/cgmlst/). A total of 2358 loci were selected and alleles were called using the chewBBACA algorithm (https://github.com/B-UMMI/chewBBACA). Minimum spanning trees (MSTs) were constructed and visualized using GrapeTree (https://achtman-lab.github.io/GrapeTree/MSTree_holder.html) based on the core-genome MLST (cgMLST). Tree nodes were positioned through dynamic rendering and node style was adjusted by fine-tuning the node size and kurtosis. Nodes were coloured by the source of the isolates and node sizes were drawn proportionally to the number of isolates.^23^

### Pangenome-derived phylogeny & Population Structure

Annotated genomes (GFF3 files) generated by Bakta comprising study isolates (*n=146*) and an India representation of *K. pneumoniae* (*n=185*) were used as input to evaluate pan-genome diversity using Panaroo (https://github.com/gtonkinhill/panaroo). A Maximum likelihood tree was constructed using RAxML Next Generation (https://github.com/amkozlov/raxml-ng) by inferring best found tree and best substitution model (GTR+GAMMA; bootstrap replicates = 1000) and visualized with iTOL (https://itol.embl.de/). Phylogenetic clusters were assigned using rhierBAPS (https://github.com/gtonkinhill/rhierbaps) specifying two cluster levels with 30 initial clusters.

## Results

### Sample collection and taxonomic profiling

During the study period, 274 samples (Livestock: n=229; Hospital effluent: n=45) were collected in the city of Vellore (Southern India) and its surroundings (Suppl Fig.1). Details of sample location are given in Suppl Table 2. *E. coli* (35.5%; 315/887) was the prominent pathogen in the livestock samples followed by *Klebsiella* spp. (16.9%; 150/887), *Proteus* spp. (16.68%; 148/887), *Aeromonas* spp. (8.45%; 75/887) and *Morgenella* spp. (5.4%; 48/805) (Suppl Fig.2). From 45 samples collected from hospital sewage sites, 21 *Klebsiella* isolates (11.2%; 21/188) were identified. *Aeromonas* spp. (34.6%), *E. coli* (20.2%), *Pseudomonas* spp. (4.8%) and other NFGNB (28.2%) constituted the remaining isolates.

### Antimicrobial susceptibility testing

The antimicrobial Susceptibility of *Klebsiella* spp. isolated from clinical specimens (*n=2,398*), livestock (*n=150*) and Hospital effluent (*n=21*) is depicted in Suppl Fig. The clinical isolates were commonly resistant to the tested antimicrobials; the susceptibility percentages were: ceftazidime (38.45%, 794/2,065), cefepime (43.42%, 989/2,278), piperacillin-tazobactam (42.31%, 977/2,309), meropenem (58.25%, 1352/2,321), gentamicin (50.73%, 551/1086), amikacin (59.66%, 1374/2,303), and minocycline (63.2%, 304/481). Similarly, isolates originating from hospital effluent were commonly MDR being resistant to most tested antimicrobials. The percentage susceptibility to both ceftazidime and cefepime was (28.57%, 6/21) while 61.9% and 38.1% were resistant to carbapenems and minocycline respectively. *Klebsiella* spp. (*n=150*) isolated from livestock specimens were highly susceptible to the tested antimicrobials, susceptibility percentages were: ceftazidime (78.62%, 114/145), cefepime (78.01%, 110/141), piperacillin-tazobactam (68.06%, 98/144), meropenem (97.95%, 143/146), gentamicin (91.72%, 133/145), amikacin (88.89, 128/144) and minocycline (88.49%, 123/139) (Suppl. Fig.3).

### Whole genome sequencing

A total of 146 isolates of *Klebsiella* spp. sourced from clinical samples (n=59), livestock (n=71) and hospital effluent (n=16) were subjected to whole genome sequencing. Illumina sequencing data were assembled to draft genome sequences (5.3 – 5.8 Mb). The contig numbers of genome assemblies, obtained by WGS, ranged from 7 to 147 contigs of >500 bps/sample in *Klebsiella* isolates with N50 values of between 97,204 and 3,986,769 bp. Genome assemblies with higher contig numbers (>150) and genome size of more than 5.8 Mb were sequenced to higher depths to obtain error-free, high-quality genome sequences. Raw genome data were submitted to the European Nucleotide Archive (ENA; https://www.ebi.ac.uk/ena/browser/) under the BioProject PRJEB58136 (Suppl Table 1).

### Clonal distribution of *Klebsiella* isolates

We identified 80 unique STs including single locus variants (SLVs) which were categorized according to the source of isolation. The population structure is depicted to be a complex structure with a few STs overlapping in source of isolation while majority being restricted to a particular niche (Fig. 1). The overlapping STs such as ST29 and ST37 were shared by isolates originating from all three sources. STs derived from hospital effluents were closer to those found in clinical isolates with the exception of isolates belonging to *K. quasipneumoniae* and *K. similipneumoniae* (ST1584). Overall, isolates originating from livestock were clonally distinct from those derived from clinical/hospital effluent isolates.

**Figure 1:**
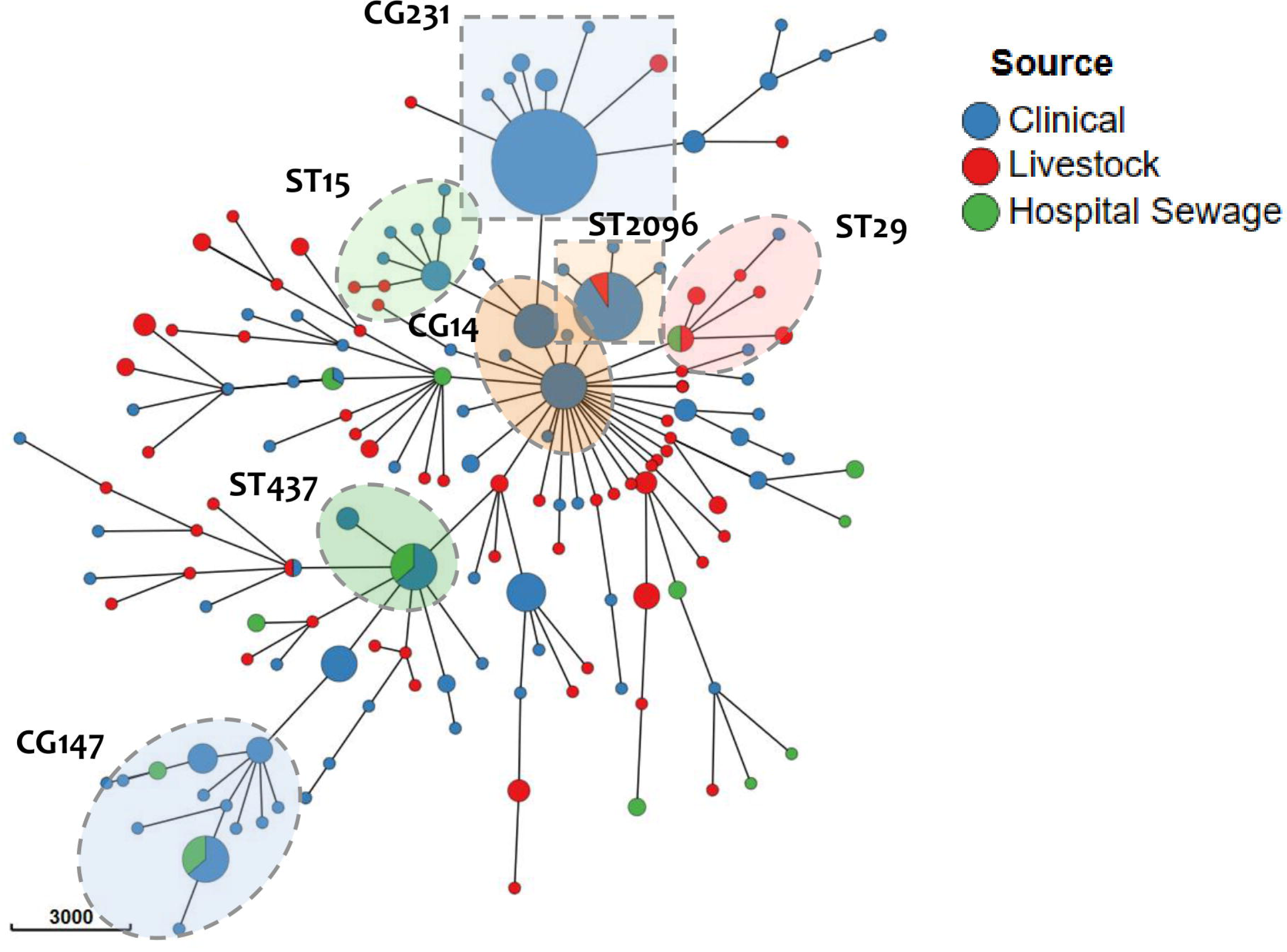
cgMLST based minimum spanning tree of *Klebsiella* spp. isolates obtained from clinical samples, livestock specimens and hospital effluent. cgMLST allele calling was performed by chewBBACA (https://github.com/B-UMMI/chewBBACA) based on the schema available in Ridom (https://www.cgmlst.org/ncs/schema/2187931/locus/). Minimum spanning tree (MSTs) was constructed and visualized using GrapeTree (https://achtman-lab.github.io/GrapeTree/MSTree_holder.html). Each node represents a unique ST or Clonal group, and node size is proportional to the number of isolates with that ST. Node color indicates the source from which the isolate was obtained. Tree nodes were positioned through dynamic rendering and node style was adjusted by fine-tuning the node size and kurtosis.

### Phylogenetic analysis

Phylogenetic analysis of 331 *Klebsiella* spp. isolates using pan-genome based core genome SNPs revealed the diversity of study isolates in the context of an Indian genomic framework. The observed core genome phylogeny clustered the isolates into two major clusters and one outgroup (Fig 2a). The outgroup consists of isolates belonging to *K. aerogenes* and *K. variicola*. The first major cluster consists of two subclusters, belonging *to K. quasipneumoniae* and *K. similipneumoniae*, and the second cluster contains isolates that have been identified as *K. pneumoniae*.

**Figure 2:**
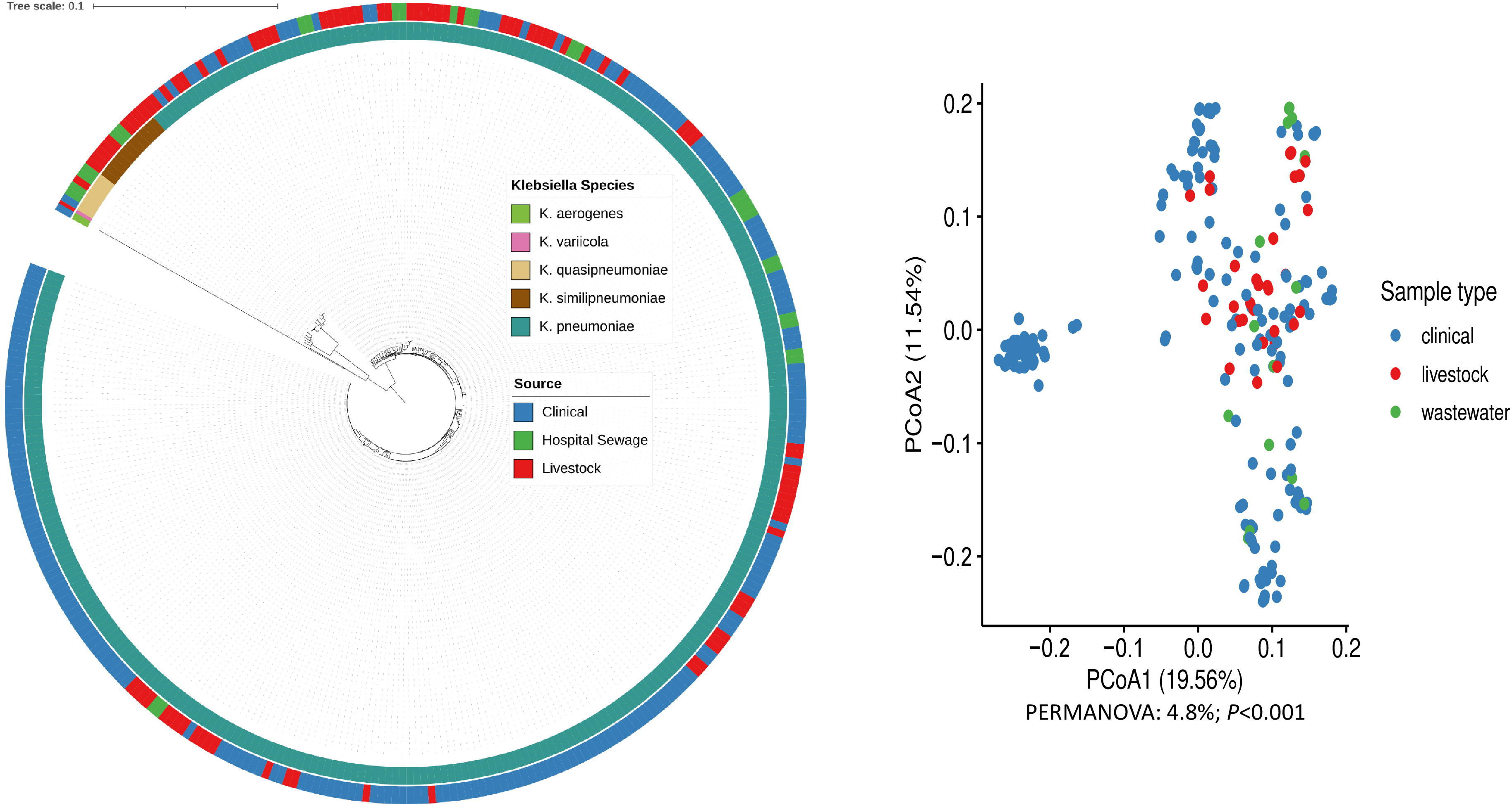
Pangenome based phylogenetic tree depicting the maximum likelihood clustering of *Klebsiella* spp based on 1,23,394 core gene SNPs. Phylogenetic tree was inferred for *n=331* genomes of *Klebsiella* spp. collected from India. Ring 1 refers to different Klebsiella species and Ring 2 represent the source of the isolates. The scale bar indicates substitutions per site. The tree was visualized and labeled using iTOL (https://itol.embl.de/). (b) Distribution of *Klebsiella* spp. isolates based on the accessory genomes.

In contrast with the MLST based clonal diversity previously observed, genome-based analyses indicate that isolates obtained from livestock and clinical settings were closely related. The isolates from different sources overlapped phylogenetically as illustrated in Fig 2a however clonal overlaps were rarely observed. There was also a high degree of overlap in the accessory genome diversity between samples from all three sources (Fig 2a). This observation suggests that the *Klebsiella* isolates collected from three different sources are clonally distinct yet genomically closely related. Overall, clinical isolates of *K. pneumoniae* were of well-defined STs while isolates originated from livestock samples were highly diverse (Fig. 2b).

### Distribution of AMR genes, virulence determinants and Plasmids

The AMR gene content of *K. pneumoniae* isolated from three distinct niches differed depending on the host. *K. pneumoniae* isolates originating from clinical specimens and hospital sewage samples generally carried a higher number of AMR genes than isolates derived from livestock samples. Clinical isolates carried at least four AMR genes (maximum 27), whereas the vast majority of livestock isolates carried only two to four acquired AMR genes. There was a wide variation in the number of carbapenem resistance determinants among isolates originating from clinical samples, hospital sewage and livestock specimens. Among the clinical samples, 76.2% (n=45/59) contained an ESBL gene, 69.5% (n=41/59) carried at least one carbapenemase gene, 71.2% (n=42/59) carried significant mutations in *ompK* genes and 6.7% (n=4/59) isolates contained a colistin resistance gene. Most isolates originating from livestock samples were predicted to be susceptible to last resort antimicrobails (ESBL: 16.9%, Carbapenamase: 2.8%, ompK mutation: 2.8%).

The Kleborate resistance score, total number of AMR genes and number of classes of AMR gene identified from isolates belonging to three different niches were represented as box plots (Fig. 3a, b & c). The clinical and hospital sewage isolates carried a median of 10 AMR genes from 6.5 classes with a resistance score of 1.5. The livestock isolates carried limited number and classes of AMR genes with a resistance score of zero (p < 0.05; Fig.3).

**Figure 3:**
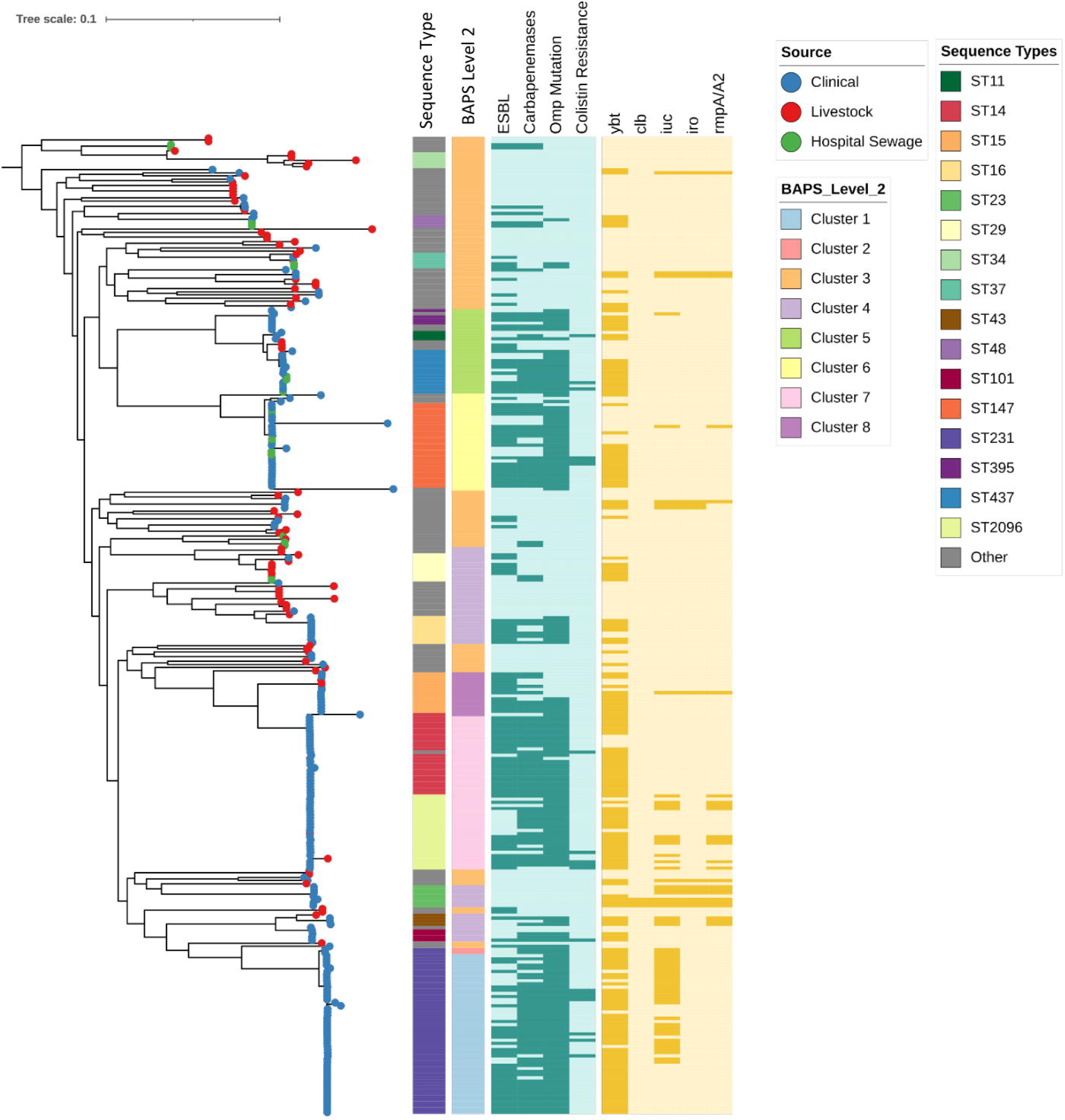
Pan-genome based phylogenetic tree of *K. pneumoniae* based on 65,505 core gene SNPs showing the comparative phylogenetic clustering by lineages, BAPS population structure, ybt-ICEKp types, K-locus types, AMR genes and virulence genes. The tree was visualized and labeled using iTOL (https://itol.embl.de/). (b) Distribution of *Klebsiella* spp. isolates based on the accessory genomes.

Comparable to previous observations, the presence of virulence genes was dependant on the sampling location. In this case both virulence scores (predicted by Kleborate) and number of virulence genes (predicted by VFDB) were significantly different for all three niches. *K. pneumoniae* isolates sourced from clinical samples showed virulence scores ranging from 1 to 4 with a median of 1 while virulence scores of isolates from hospital sewage and livestock ranged from 0 to 1 with a median score close to 0 (Fig. 4). The total number of virulence genes also showed similar variation (Clinical > Hospital sewage > Livestock). This observation suggests that hyper-virulent clones of *K. pneumoniae* isolates are commly found in clinical samples and not any other niches.

**Figure 4:**
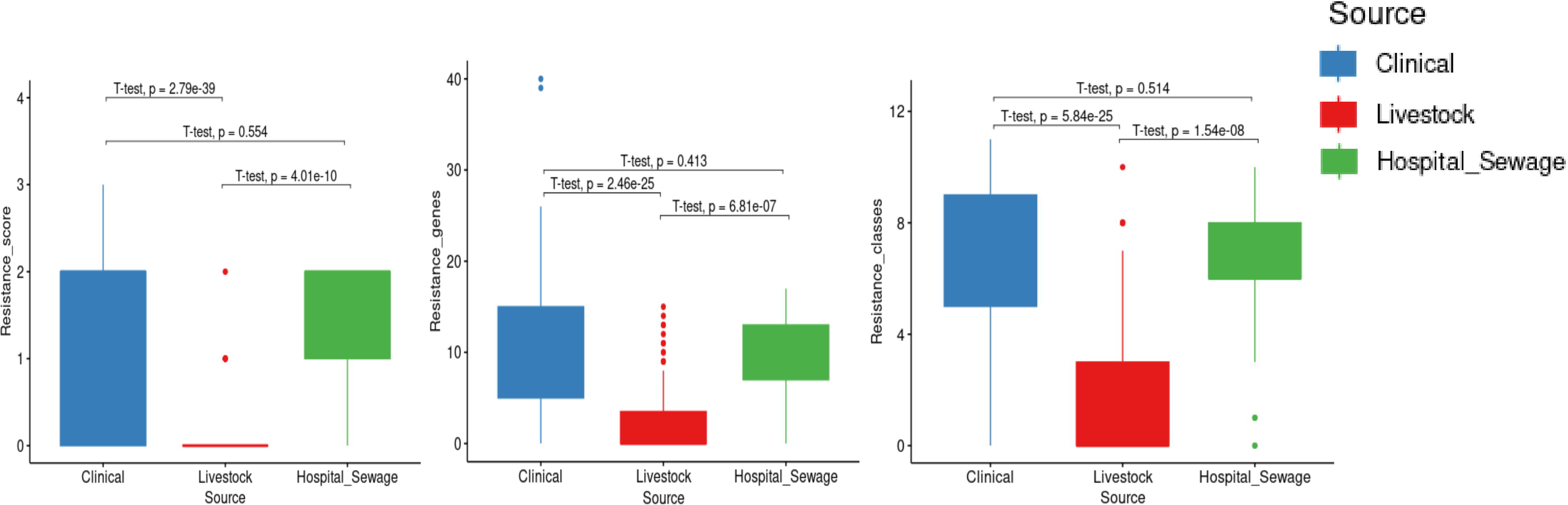
(a) Resistance score of *K. pneumoniae* isolates sourced from clinical, livestock and hospital sewage samples as predicted by Kleborate (b) Total number of antimicrobial resistance genes identified in *K. pneumoniae* isolates sourced from clinical, livestock and hospital sewage samples (c) Total number of classes of antimicrobial resistance genes identified in *K. pneumoniae* isolates sourced from clinical, livestock and hospital sewage samples.

Plasmid replicons carried by *K. pneumoniae* isolates were also found to be host specific. *K. pneumoniae* isolates originated from clinical and hospital sewage samples exhibited 1 to 8 plasmids with a median of 5 plasmids while isolates derived from livestock samples carried 0 to 6 with a median of 2.5 plasmids (Fig. 5). In clinical and hospital sewage samples, there were significantly more plasmid replicons than in livestock samples (p=1.191x 10^-19,^ p < 0.05; Fig.4).

**Figure 5:**
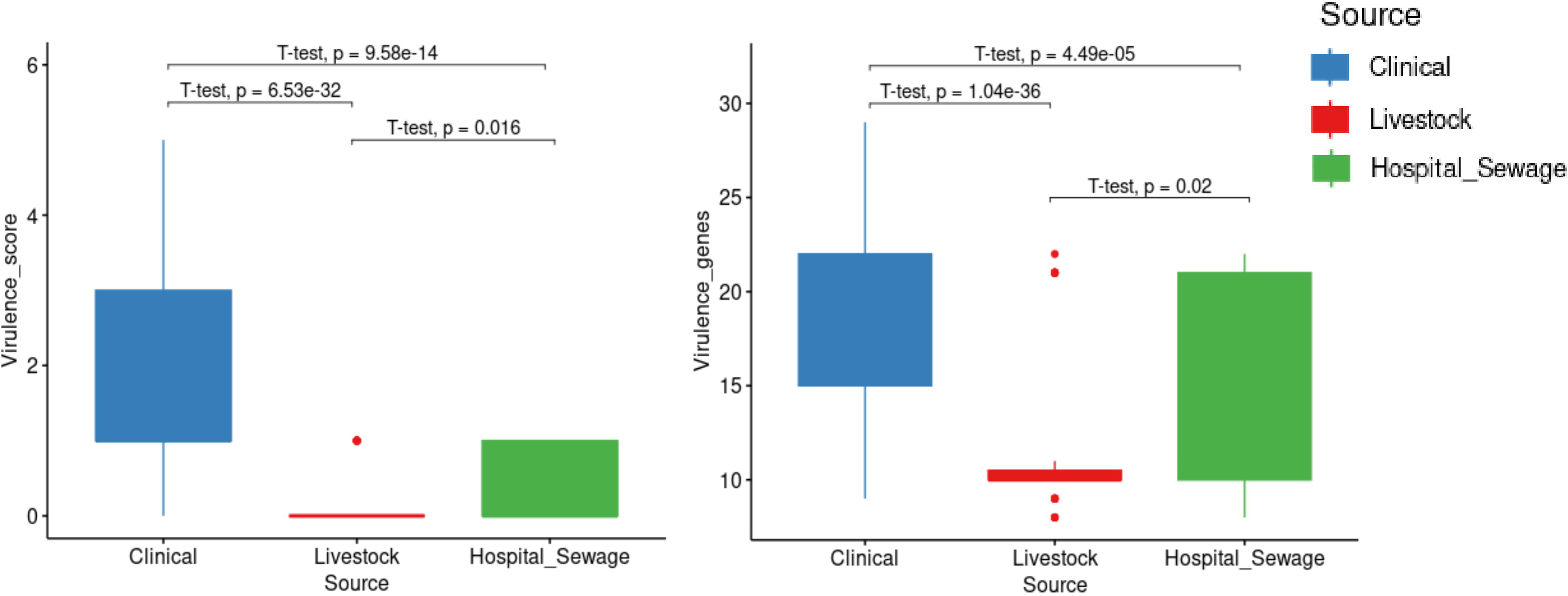
(a) Virulence score of score of *K. pneumoniae* isolates sourced from clinical, livestock and hospital sewage samples as predicted by Kleborate **(b)** Total number of virulence genes identified in *K. pneumoniae* isolates sourced from clinical, livestock and hospital sewage sample.

## Discussion

The emergence of AMR contributes significantly to increasing morbidity and mortality. MDR/XDR *K. pneumoniae* is the major cause of difficult-to-treat infections.^24, 25^ Increased isolation rates of carbapenem-resistant *K. pneumoniae* in hospital settings have often been attributed to inadequate infection control practices.^26, 27^ In addition, wide range of ecological niches, such as food animals, wastewater, and plant-based foods have also been identified as reservoirs and transmission sources of AMR.^8^ AMR transmission routes may therefore be studied using *K. pneumoniae* due to its high genome plasticity and AMR trafficking capabilities.^8, 25^ Using the One Health framework, we have been able to integrate humans, animals, and other environmental reservoirs of AMR, thereby establishing a multisectoral approach to investigate AMR.

There is a well-established pattern of prevalence and distribution of clones, resistance, and virulence determinants among clinical strains of *K. pneumoniae* in India.^28, 29^ Several previous studies have demonstrated that the proportion of carbapenemase-producing isolates varies by location and study, ranging from 22% to 66%.^27, 30, 31^ In contrast, characterisation of *K. pneumoniae* from non-clinical sources has been poorly documented apart form a few small localized studies.^32, 33^ In the present study we detected phenotypic carbapenem resistance in 41.75% of clinical isolates during 2020 - 2022. This is slightly higher than the previous report (32%) from our hospital in 2016 for CR-Kp.^27^ The analysis of *K. pneumoniae* isolates derived from non-clinical sources indicated that a larger proportion of carbapenem-susceptible isolates were recovered from livestock (98%) than hospital sewage (62%). This is in accordance with recent studies that showed most hospital-associated strains were resistant to last resort antimicrobials such as carbapenems, while livestock associated isolates were not. ^13, 14, 34, 35^

We compared the clonal complexes of *K. pneumoniae* across three different niches to understand the distribution of these systematically collected isolates. The clonal distributions of *K. pneumoniae* were generally diverse within and between the niches (human, livestock, and hospital effluent samples). The diversity across the niches can be attributed to host-specific or environment-specific drivers.^8^ In line with our earlier findings, ST231, CC14, CC147, and ST2096 remain the most prevalent clones among clinical isolates.^28^ Except for CC147 from hospital sewage, none of the other high-risk *K. pneumoniae* clones have been isolated from non-clinical samples. Further, several prominent clones found in livestock, including ST29, ST138, and ST2735, are rarely reported to cause nosocomial infections.^36, 37^ The overlapping sequence types between clinical and livestock samples, such as ST2096, ST29, ST37 and ST152, was also observed sporadically among livestock isolates (Fig. 6). Because *K. pneumoniae* clones within hospital settings differ significantly from those found in livestock samples, it is difficult to verify reports that suggesting high-risk clones emerged outside of hospitals.

**Figure 6:**
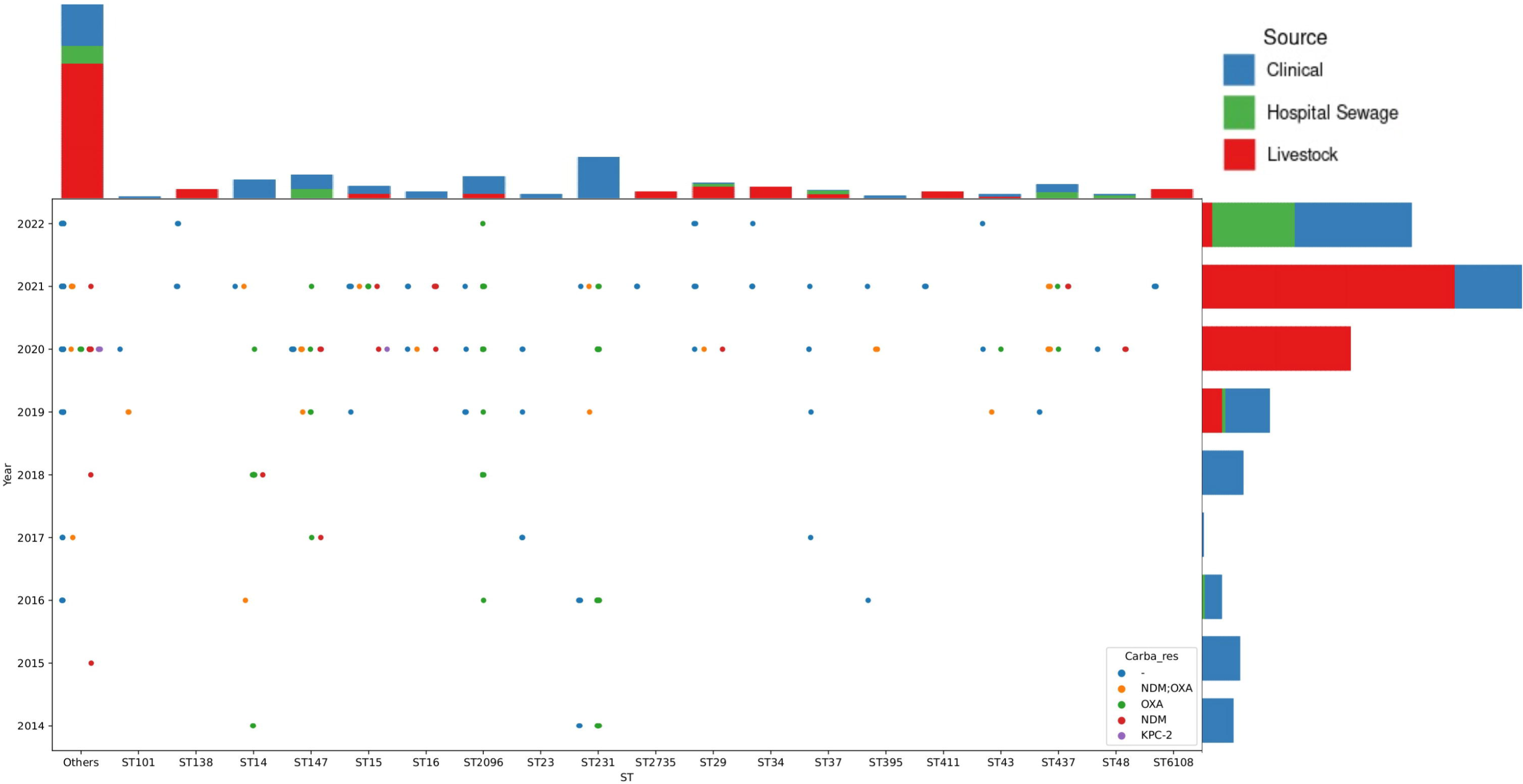
MLST, source and carabapenamase gene distribution of *K. pneumoniae* isolates sourced from clinical, livestock and hospital sewage samples as predicted by Kleborate

Phylogenetic analysis confirmed the classification of *K. pneumoniae* species complex (KpSC) into five predefined phylogroups, with *K. pneumoniae* (Kp1) being the dominant group.^38^ Though the clonal group-wise organization of *K. pneumoniae* is clearly compartmentalized, the lineages were not separated according to ecological niche. The distribution of isolates within phylogroups were consistent with the STs but again not according to the niche adaptation discussed earlier. This suggests that the niche adaptation is yet to promote evolutionary diversification in *K. pneumoniae* or the adaptive perspective is not clear from the genome sequencing data.^39^ Phylogenetic comparisons of *K. pneumoniae* by taking account of clonal complexes illustrated that most non-clinical isolates were not closely related to high-risk clones ST231, ST2096 and CC147. When considering the possibility of clonal spread, it is more likely that it will occur within settings (hospital outbreaks) than between them.^40^

Our analysis also indicated increased prevalence of AMR genes, virulence determinants and mobile genetic elements in both clinical and hospital effluent isolates. Moreover, most of the hospital-associated clones possess resistance genes against last-resort antimicrobials such as 3^rd^/4^th^ generation cephalosporins, carbapenems, or minocycline. As reported previously, livestock isolates exhibited less abundance and diversity of acquired AMR genes, and the mechanisms of resistance were primarily directed against first-line drugs.^41,42^ Based on these findings, it appears that AMR prevalence in different niches depends on a variety of site-specific conditions, such as antimicrobial use intensity and residual concentrations.^43^ Similarly, livestock isolates contained significantly fewer virulence genes and mobile genetic elements than hospital-associated isolates. Isolates containing hypervirulent markers, such as *iuc, iro, rmpA*, and *rmpA2*, were rare, and these genes are mostly found in convergent nosocomial clones.^44^ Isolates belonging to these clones belong to carbapenem resistant - hypervirulent - high-risk clones, namely ST231 and ST2096.^45^ Considering that *K. pneumoniae* have a low fitness burden for plasmid carriage and maintenance, the absence of resistance plasmids in livestock isolates is rather surprising.^8^ These data suggest the possibility of specific adaptations of *K. pneumoniae* clones to change the plasmid permissiveness. It is clear that AMR genes and plasmids are not randomly distributed amongst *K. pneumoniae* lineage instead associated to specific environments based on adaptations.^8^

The limited overlap of *K. pneumoniae* clones, AMR genes, virulence determinants, and plasmids across different settings opposes the current perception of AMR transmission.^10^ In line with the recent finding, this data challenges the current view of the One-Health approach at least in terms of AMR transmission.^14, 40^ Accordingly, we found that the adaptation of K. pneumoniae to local environments plays an important role in its evolution and therefore in mitigating the spread of AMR. The distribution of *K. pneumoniae* in different settings are not randomly distributed, but rather specialized for particular host environment.^46^ At present, vast majority of MDR/XDR *K. pneumoniae* in humans is currently acquired from other humans, which is also consistent with previous studies.^14, 40^ Since there is no evidence of highly virulent or resistant isolates of *K. pneumoniae* outside of clinical settings, it may be possible for barriers to exist between different settings.

There are several limitations to our study. We acknowledge that we sequenced only a representative collection of 146 isolates that were collected during the study period. The selection of isolates for sequencing based on antibiogram could potentially introduce bias into the study. Secondly, our sampling strategy may not have adequately capture the true distribution of Klebsiella isolates across different sources. Most importantly, there is a lack of data about antimicrobial use in samples of food animal collected for this study. Moreover, *K pneumoniae* isolated from humans are only represent by hospital-acquired isolates. A more accurate prediction of antimicrobial resistance transmission dynamics would have been achieved by including isolates representing community-acquired infections. Hence, additional studies are needed to determine whether our findings will be replicated in other study settings.

## Conclusion

*K. pneumoniae* exhibits a high level of resistance and virulence in comparison with other *Klebsiella* species. The spread of XDR or hypervirulent clones of *K. pneumoniae* appear to be confined to humans and no clear linkage with non-clinical sources could be established in this setting. In general, our findings suggest that hospitals are considered hotspots of dissemination of *K. pneumoniae* resistance in India, while isolates originating from livestock samples are largely susceptible to current antimicrobial therapies. Moreover, the currently emerging convergent clones of *K. pneumoniae* carrying both resistance and virulence determinants (ST231, ST2096) are likely to have been emerged in hospital settings rather than in animal or natural environments. The One Health approach to AMR transmission continues to be relevant for sporadic transmission events, and transmission dynamics may differ depending on the region and pathogen being studied.

## Data Availability

Raw read data were deposited in the European Nucleotide Archive (ENA) under project accession number: PRJEB58136. The individual sample accession numbers are listed in Supplementary Table

https://www.ncbi.nlm.nih.gov/bioproject/PRJEB58136/

---

**Suppl Fig. 1:** Map showing the location of sampling and study areas in Vellore District, Tamil Nadu, India during 2020 – 2022. The map was exported and processed using QGIS software version 3.32 (https://www.qgis.org/en/site/).

**Suppl Fig. 2:** Taxonomic distribution of bacterial isolates identified from Livestock samples

**Suppl Fig 3:** Antimicrobial Susceptibility of *Klebsiella* spp. isolated from clinical specimens, livestock samples and hospital effluent. Percent susceptibility displayed reflects the percentage of susceptible isolates among all isolates with antimicrobial susceptibility testing data is available. Percent susceptibility is not displayed in cases where more than 50% of isolates were not tested for susceptibility to a given antibiotic.

## Acknowledgements

We would like to express our gratitude to Dr. John Antony Jude Prakash Head of Clinical Microbiology, CMC Vellore for his support and guidance. We would like to thank Dr. Solomon D’Cruz for his assistance in delineating the sampling sites and locating them on the base map using QGIS. Many Thanks to the support staff of the Department of Clinical Microbiology CMC Vellore.

## Ethical Clearance

Institutional Review Board (IRB) of Christian Medical College (CMC), Vellore, India gave ethical approval for this work vide IRB Min no. 12626 dated 26.02.2020. As the required data had been collected as part of the standard of care for diagnosis, informed consent waiver was granted by the IRB committee CMC, Vellore.

## Contributors

Conceptualization: BV and KW

Methodology and Investigation: JJJ, AV, BSBJ, BLY and CS

Data analysis: JJJ and AV

Visualization: JJJ, AV and BSBJ

Funding acquisition: BV and KW

Project administration: JJJ, CS, YM, FA, MPT, SK

Supervision: BLY, BV and KW

Original draft: JJJ, AV, BSBJ: Writing

Writing – Review & editing: SB, BV and KW

## Disclaimer

The funders had no role in the design and conduct of the study; collection, management, analysis, and interpretation of the data; preparation, review, or approval of the manuscript; and decision to submit the manuscript for publication.

## Patient consent for publication

Not required.

## Data availability statement

Raw read data were deposited in the European Nucleotide Archive (ENA) under project accession number: PRJEB58136. The individual sample accession numbers are listed in Supplementary Table 1

