## Supplementary figures and images for "Limited evidence of spill over of antimicrobial resistant *Klebsiella pneumoniae* from animal/environmental reservoirs to humans in India"

### supplementary Fig 1

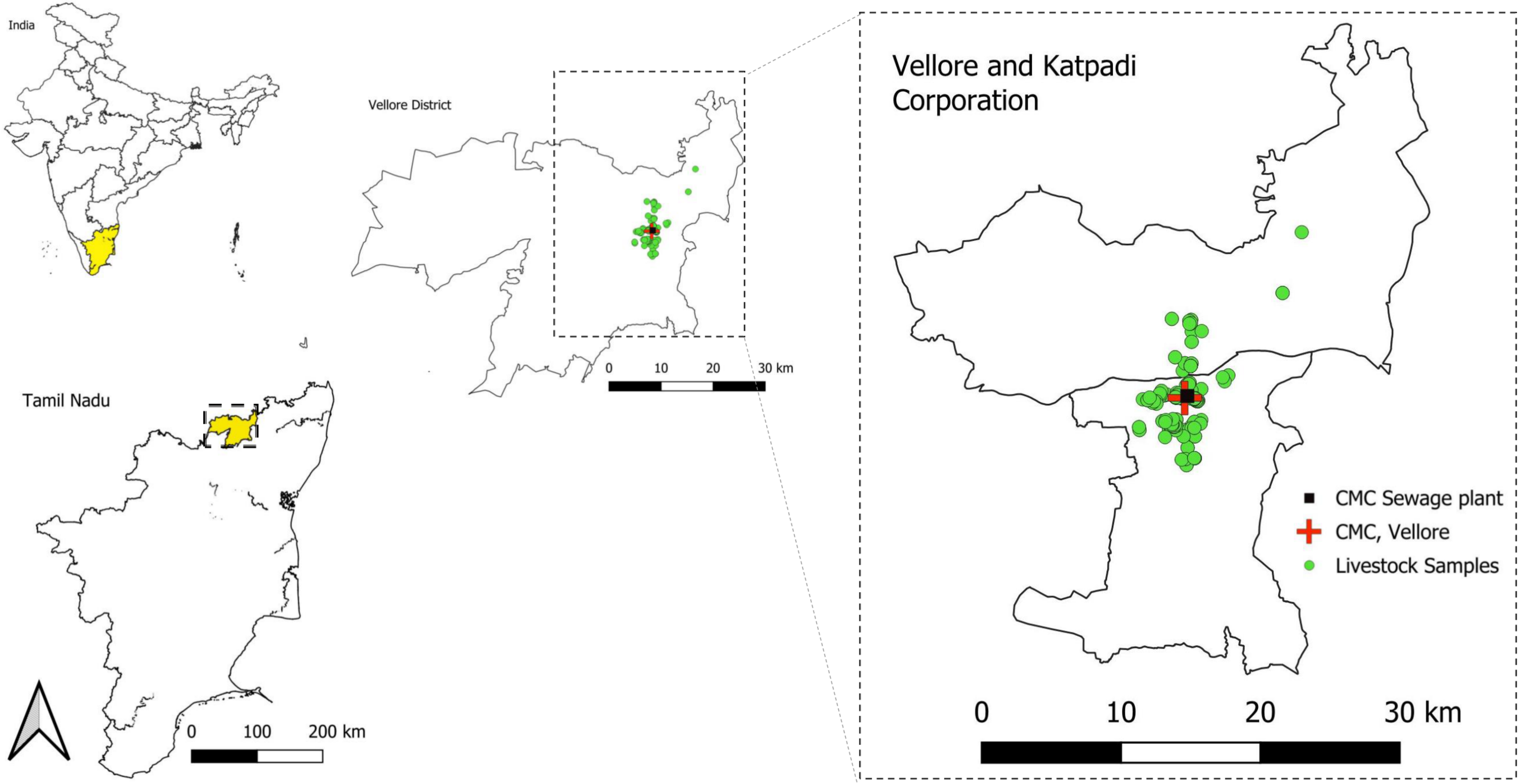

### supplementary Fig 2

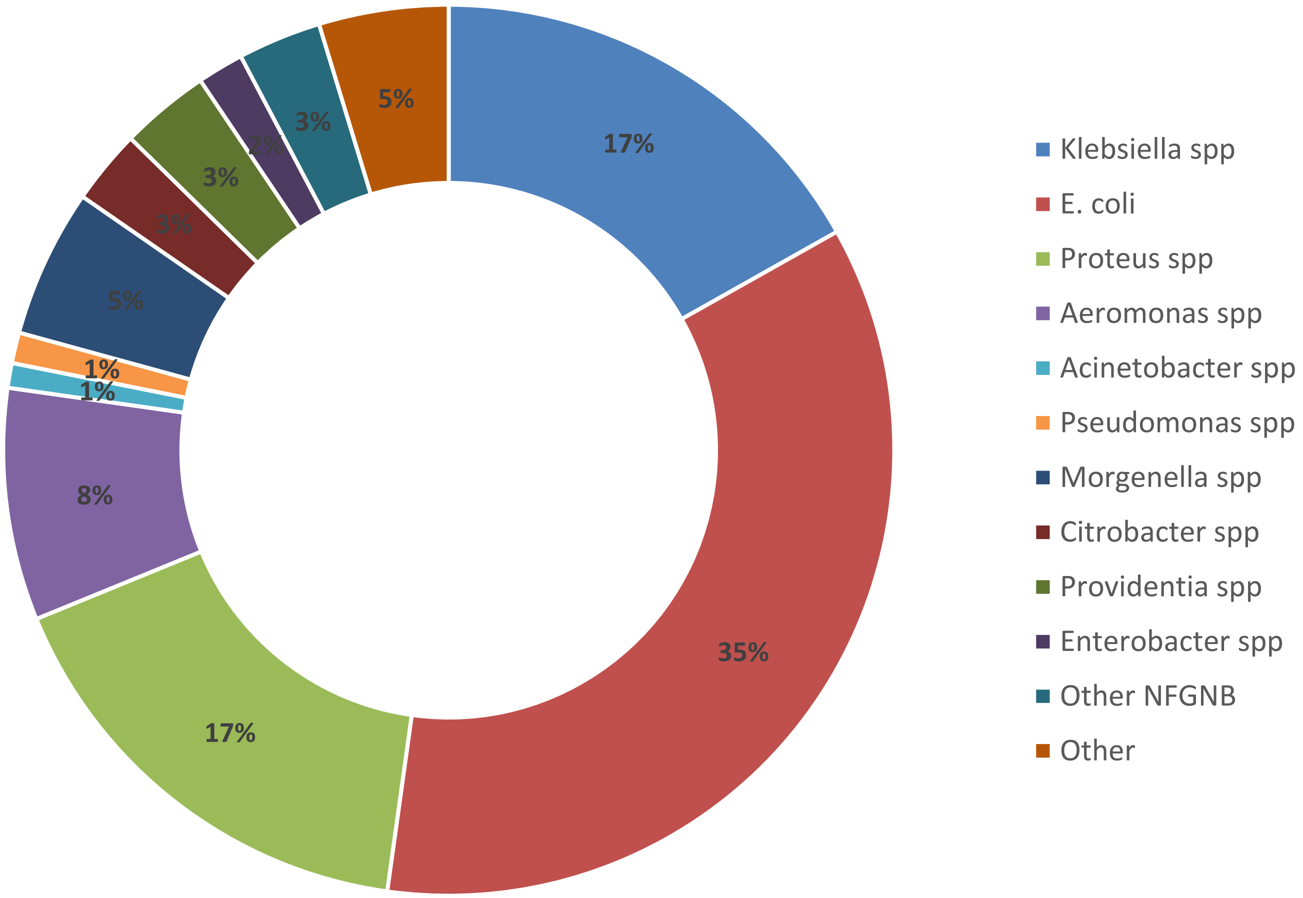

### supplementary Fig 2

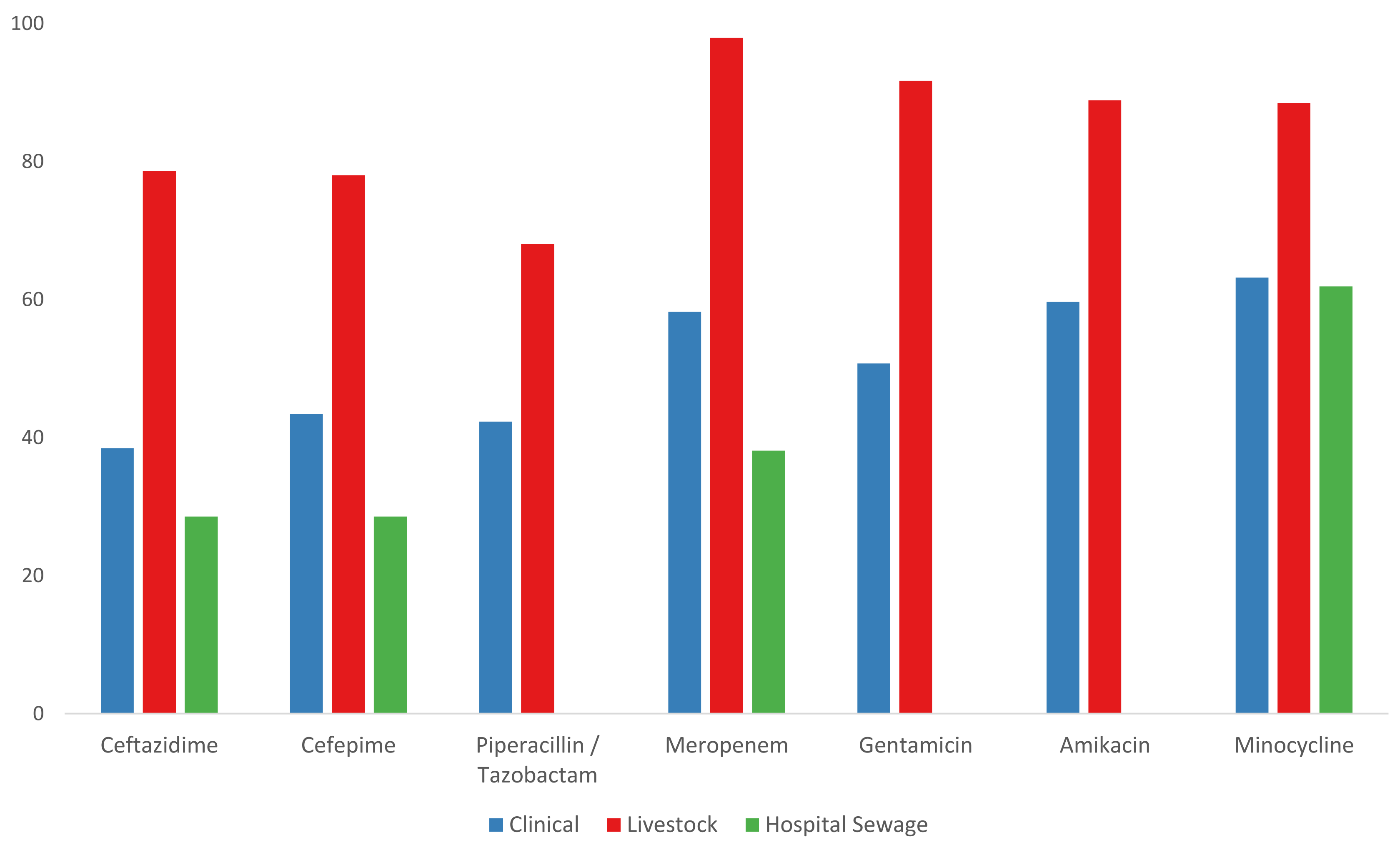
